# Mapping echocardiogram reports to a structured ontology: a task for statistical machine learning or large language models?

**DOI:** 10.1101/2024.02.20.24302419

**Authors:** Suganya Subramaniam, Sara Rizvi, Ramya Ramesh, Vibhor Sehgal, Brinda Gurusamy, Hikamtullah Arif, Jeffrey Tran, Ritu Thamman, Emeka Anyanwu, Ronald Mastouri, G. Burkhard Mackensen, Rima Arnaout

**Affiliations:** University of California, San Francisco, San Francisco, California; University of California, Berkeley, Berkeley, California; University of Washington, Seattle, Washington; University of Arizona, Tucson, Arizona; University of Pittsburgh, Pittsburgh, Pennsylvania; University of Pennsylvania, Philadelphia, Pennsylvania; Indiana University, Indianapolis, Indiana

**Keywords:** natural language processing, machine learning, large language models, echocardiography report, ontology

## Abstract

**Background:** Big data has the potential to revolutionize echocardiography by enabling novel research and rigorous, scalable quality improvement. Text reports are a critical part of such analyses, and ontology is a key strategy for promoting interoperability of heterogeneous data through consistent tagging. Currently, echocardiogram reports include both structured and free text and vary across institutions, hampering attempts to mine text for useful insights. Natural language processing (NLP) can help and includes both non-deep learning and deep-learning (e.g., large language model, or LLM) based techniques. Challenges to date in using echo text with LLMs include small corpus size, domain-specific language, and high need for accuracy and clinical meaning in model results.

**Methods:** We tested whether we could map echocardiography text to a structured, three-level hierarchical ontology using NLP. We used two methods: statistical machine learning (EchoMap) and one-shot inference using the Generative Pre-trained Transformer (GPT) large language model. We tested against eight datasets from 24 different institutions and compared both methods against clinician-scored ground truth.

**Results:** Despite all adhering to clinical guidelines, there were notable differences by institution in what information was included in data dictionaries for structured reporting. EchoMap performed best in mapping test set sentences to the ontology, with validation accuracy of 98% for the first level of the ontology, 93% for the first and second level, and 79% for the first, second, and third levels. EchoMap retained good performance across external test datasets and displayed the ability to extrapolate to examples not initially included in training. EchoMap’s accuracy was comparable to one-shot GPT at the first level of the ontology and outperformed GPT at second and third levels.

**Conclusions:** We show that statistical machine learning can achieve good performance on text mapping tasks and may be especially useful for small, specialized text datasets. Furthermore, this work highlights the utility of a high-resolution, standardized cardiac ontology to harmonize reports across institutions.

## Introduction

Big data has the potential to revolutionize echocardiography by enabling novel research and rigorous, scalable quality improvement^1^. Echocardiogram text reports are a key component of such analyses, serving as the prime means of communication for imaging findings ^2^ and as a source of data labels for machine learning research.

Currently, echocardiogram reports include both structured and free text and vary across institutions, hampering attempts to mine text for useful insights. Alternatively, mapping report text to a standardized ontology can help harmonize reports across institutions, languages, and imaging modalities^3^.

Several medical ontologies exist ^4–7^. The Unified Medical Language System (UMLS)^4^ is supported in some natural language processing (NLP) software packages, but it is focused on describing terms found in clinical notes rather than the structure and attribute details important in echocardiogram reporting. Radlex^6^, developed and maintained by the Radiological Society of North America (RSNA), is not as well supported by NLP software but contains additional attributes, for example on patient status and study protocol, that may be useful for echocardiography as well. To date, however, neither echocardiogram nor radiology report text is routinely mapped to an ontology in clinical practice.

Similarly, NLP research to date has focused not on mapping text to ontology but on extracting particular numerical values or concepts of interest from medical report text. Many published methods for extraction of multiple values use tailored regular expressions and rule- or pattern-based algorithms^8–12^. These approaches are an effective use of small datasets, but they can require considerable human effort to develop and can be poorly generalizable to additional concepts or institutions outside the training set. Most did not test externally^9–12^, and one that did showed recall near 50% on the external test set^8^.

Machine learning has been used on clinical reports but not specifically for extraction of concepts from echocardiogram reports. Many have used various implementations of BERT (Bidirectional Encoder Representations from Transformers), an early large language model (LLM), to extract radiographic clinical findings^13^, mentions of devices^14^, study characteristics^15^, and result keywords^16^ from radiology or pathology reports.

However, performance of these models is often lower than what would be required clinically without additional feature engineering^13,15^ or fine-tuning on thousands of manually-derived labels^14,16^ specific to the task. This likely reflects the fact that medical report text has specific structure and meaning while comprising only a small proportion of the general language used to train these models. Newer, larger LLMs like GPT hold greater promise for one-shot inference^17,18^ (making predictions without the need for transfer learning or fine-tuning) so that all curated data may be used for testing. To our knowledge these models have not been applied specifically to echocardiogram report text.

In this work, we use NLP to extract all qualitative components of an adult transthoracic echocardiogram (TTE) report and map them to a standardized hierarchical ontology in a way that accommodates both free and structured text from across institutions. We compare a statistical ML model we developed, EchoMap, against one-shot inference using GPT.

## Methods

### Ontology construction

We developed a three-tier ontology for the echocardiographic anatomic structures, functional elements, and descriptive characteristics in adult transthoracic echocardiograms. UCSF’s structured echocardiogram report data dictionary (standard phrases that populate structured reporting, n=951 sentences) was used as a starting point. Structures or attributes not mentioned in UCSF dictionaries were added, and all terms were mapped to UMLS (https://uts.nlm.nih.gov/uts/umls/home) and RadLex (https://radlex.org/) identifiers where available.There were 260 distinct ontology terms across all three levels of the ontology (five terms are found in both Level 2 and Level 3 of the ontology). Table 1 provides an example.

**Table 1.** Example sentences from TTE reports and representative ontology mappings.

|  | In TTE Ontology |  | Not in TTE Ontology |
| --- | --- | --- | --- |
|  | In UMLS/Radlex | Not in UMLS/Radlex |  |
| **Example Sentence** | LV systolic function appears hyperdynamic. | Small VSD is seen in the perimembranous septum. | There is no aortic valve mass present. |
| **L1** | LEFT VENTRICLE | INTERVENTRICULAR SEPTUM | AORTIC VALVE |
| **L2** | FUNCTION | DEFECT | MASS |
| **L3** | HYPERDYNAMIC | SMALL PERIMEMBRANOUS | NONE |

### Datasets

<u>Training and validation.</u> The UCSF data dictionary (n=951 sentences) was split into a training set (n=723 sentences) and separate validation set (n=228 sentences).

<u>Testing.</u> Data dictionaries from the University of Arizona (n=1202), Indiana University (n=2143), University of Washington (n=504), University of Pennsylvania (n=2024), and University of Pittsburgh (n=966) medical centers were used as five external test sets. Two additional test sets were derived from patient echocardiogram reports. First, all patient echo reports from 1995-2021 were obtained from UCSF in accordance with the UCSF IRB and de-identified. 102 reports in the UCSF system had come from eighteen outside hospitals and were used for an “outside hospital” dataset (n=483 unique sentences). Second, UCSF patient reports had data dictionary sentences removed so that only free-text sentences were remaining (n >250,000 sentences). A random sample of these free-text sentences (1500 minus 32 sentence fragments removed, n = 1468 unique sentences) became the UCSF free-text dataset.

All training and test datasets were labeled with ontology terms by clinicians, to serve as ground truth.

### Text processing

Sentences were delimited, spell-corrected, abbreviations were expanded, and made lowercase.

### Feature engineering

For input into the statistical ML model, feature engineering was performed on each sentence as follows. For each sentence, negation detection (https://pypi.org/project/negspacy/) was performed to tag sentences with negation detection absent vs present (0 vs 1). UMLS entities were extracted from each sentence (https://pypi.org/project/scispacy/ named entity recognition (NER), and principal component analysis (PCA) (https://scikit-learn.org/) was used to reduce this result into a 25-element vector. Finally, Jaccard indices (a measure of overlap) were calculated between the sentence and each ontology term (257 terms) using tri-grams. The final input feature to the Level 1 statistical model was 53 elements (negation + 25-element UMLS + 27 Level1 Jaccard indices). The final input feature to the Level 2 statistical model was 284 elements (negation + UMLS + 27 Level1 Jaccard + 230 Levels 2 and 3 Jaccards + Level1 prediction). The final input feature to the Level 3 statistical model was 285 elements (Level2 features + Level2 prediction).

Additional features calculated, but not used in the final model, include a 25-element embedding from position of speech tagging (“en_core_web_sm” model from scispacy, then reduced to 25 elements with PCA) and a 25-element embedding from the BioWordVec large language model^19^ (“BioWordVec_PubMed_MIMICIII_d200.vec.bin” from scispacy, reduced with PCA (Supplemental Table 2).

For inference using GPT 3.5 (OpenAI), the OpenAI API was accessed using scikit-llm (https://pypi.org/project/scikit-llm/ ). For input into GPT3.5, raw sentences were used without feature engineering.

### Model architectures, training, and inference

For the statistical ML model, random forest (RF) classifiers were used (https://scikit-learn.org/) for classification at each of the three ontology levels. GridSearch was used to help determine optimal parameters for each classifier. The first RF classifier had a maximum depth of 25 and 100 estimators. The second classifier had max depth 40, number of estimators 120. The third classifier had max depth 25, number of estimators 100. All classifiers used the class balance option to mitigate class imbalance among different ontology terms.

For the large language model (LLM) classifier, OpenAI’s GPT 3.5 (a pre-trained model) was accessed via scikit-llm Python package (https://pypi.org/project/scikit-llm/; the zero-shot GPT classifier and multi-class zero-shot GPT classifiers were used, respectively, for one classification at a time vs multi-class predictions).

### Statistical analysis

Model predictions were compared to ground-truth labels, and percent correct was calculated both by ontology level and overall. One-sample t-test, Friedman *X*^2^, and Nemenyi post-hoc comparisons were used to perform statistical comparison across groups.

### Data and code availability

Code will be made available at github.com/ArnaoutLabUCSF/CardioML/ upon publication. Patient report data cannot be made available. The UCSF data dictionary can be made available upon reasonable request for non-commercial use and with approval.

## Results

An ontology was created from the UCSF data dictionary as a proof-of-concept ontology designed to capture most relevant TTE descriptors within three or fewer hierarchical levels (Table 1). We then tested the ability of (i) a hierarchical statistical machine learning model, termed “EchoMap,” trained on a small corpus of echocardiogram sentences and (ii) one-shot inference from GPT to map echocardiogram report sentences to the ontology. Overall, EchoMap outperformed single-shot GPT.

### Structured reporting text varies across institutions, and existing medical ontologies do not contain all terms relevant to echocardiography

Despite all adhering to clinical guidelines, there were notable differences by institution in what structural and functional information was included in structured reporting. Our proof-of-concept TTE ontology (that captures all concepts in the UCSF data dictionary) captured only 57-68% of concepts in the data dictionaries from the other institutions in our test set (Table 2).

**Table 2.**
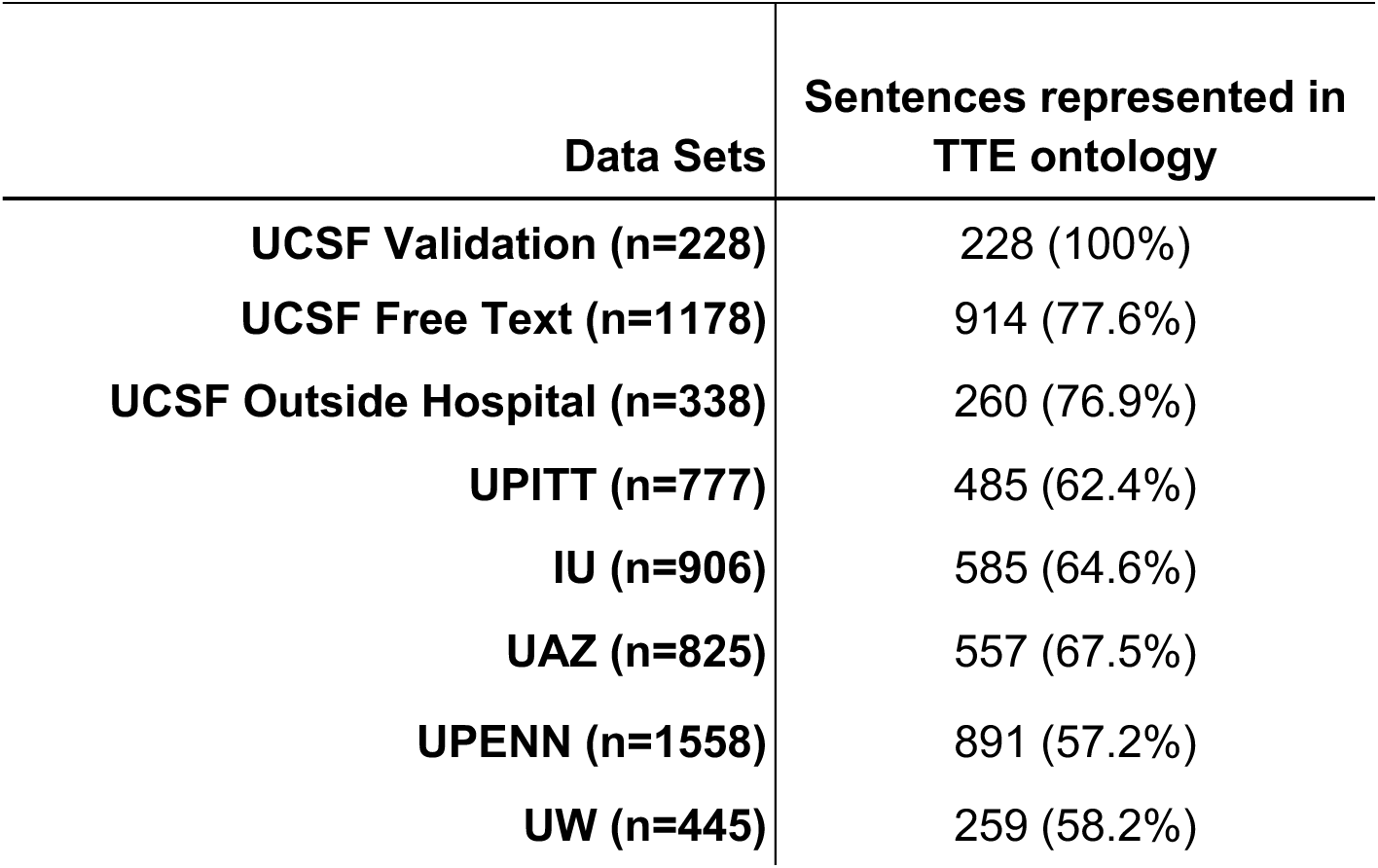
Sentences in external data dictionaries represented by TTE ontology.

Additionally, mainstream ontologies like UMLS and Radlex contained some, but not all, of the echo-specific terms from the TTE ontology (Table 3). At each level of our ontology, UMLS contained a higher proportion of terms than Radlex did (while Radlex includes additional terms describing radiology study quality and protocol, those are not part of the current TTE ontology). Level 1 of the ontology, containing more common cardiac structures, was best covered in both UMLS and Radlex, followed by Level 2, and then Level 3, which each contain structures and observations successively more specific to echocardiogram findings.

**Table 3.** TTE ontology concepts represented in existing ontologies.

|  | **UMLS** | **Radlex** |
| --- | --- | --- |
| **Overall (n=260)\*** | 189 (72.7%) | 84 (32%) |
| **UCSF Ontology Level** |  |  |
| Level 1 (n=27) | 24 (89%) | 18 (67%) |
| Level 2 (n=102) | 88 (86%) | 39 (38%) |
| Level 3 (n=150) | 95 (63%) | 39 (26%) |

### Machine learning can map echocardiography report text to an ontology

We tested two machine learning models for mapping echocardiogram report sentences to our TTE ontology. The first was EchoMap, a statistical machine learning model trained on a portion of the UCSF data dictionary (see Methods). The second was single-shot inference using GPT. We used GPT in two ways: (i) single-shot inference to predict all three ontology terms at once (multi-class inference), and (ii) single-shot inference to predict Level 1, Level 2, and Level 3 terms separately (Figure 1).

**Figure 1.**
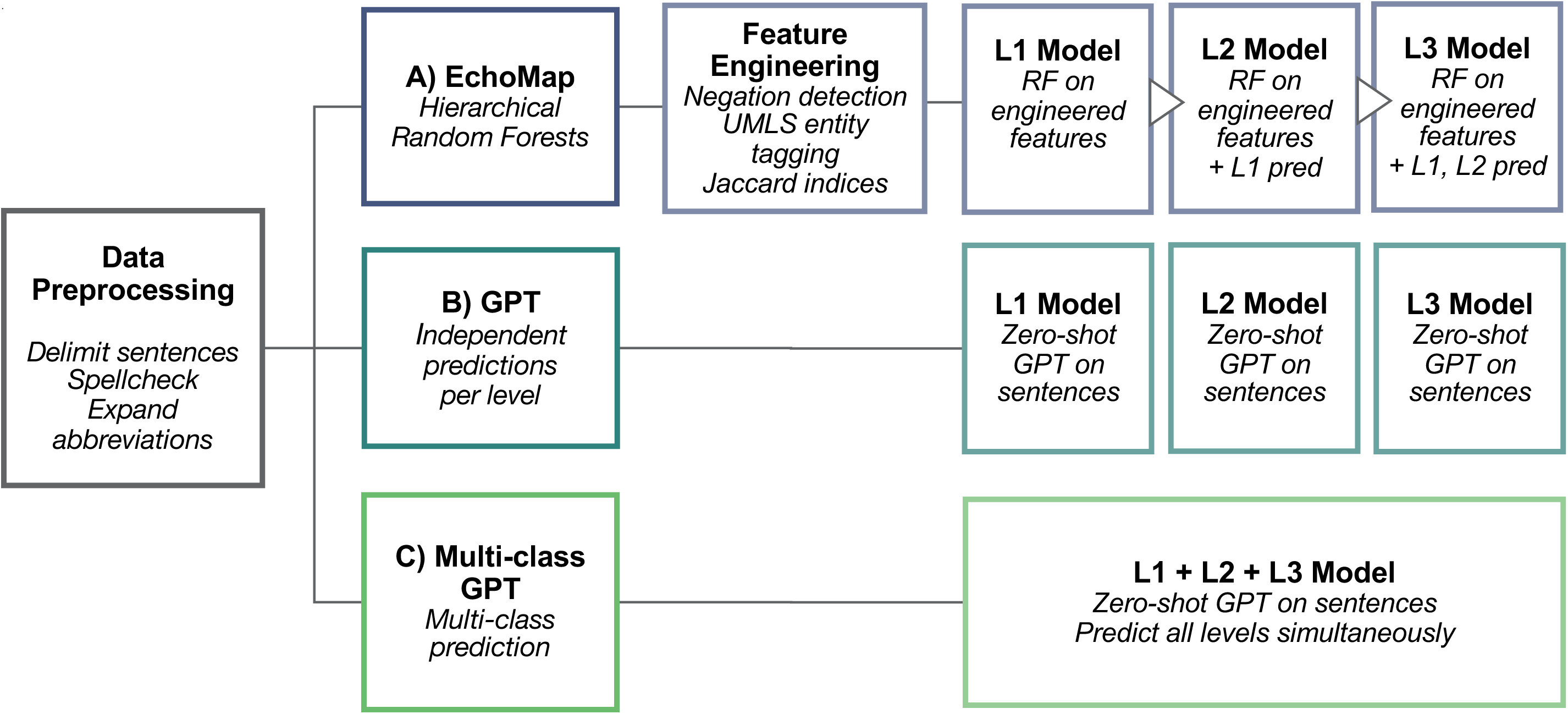
Workflow for the three machine learning approaches evaluated. Data (structured dictionaries and free text from echo reports) were preprocessed, then passed to each of three model types: **(A)** Hierarchical Random Forest statistical machine learning model, which included additional engineered features and used each level’s prediction to inform the subsequent level, **(B)** Zero-shot GPT making independent predictions per level of ontology, **(C)** Zero-shot GPT making multi-class prediction. GPT, Generative Pre-trained Transformer. RF, Random Forest. UMLS, Unified Medical Language System. L1, L2, L3, Level 1, Level 2, Level 3, respectively.

For EchoMap, the balance of the UCSF dictionary served as a validation dataset, and seven additional datasets served as test datasets. These included the data dictionaries from the University of Washington, the University of Pennsylvania, the University of Pittsburgh, Indiana University, and the University of Arizona, as well as free text sentences from UCSF reports, and report sentences from a group of 18 outside hospitals incidentally found in the UCSF database. Because GPT was used “out of the box” rather than fine-tuned on any of our echocardiogram text, the UCSF data dictionary also served as a test set with respect to GPT.

EchoMap’s validation accuracy was 98% for the first level of the ontology, 93% for the first and second levels together, and 79% for the first, second, and third levels (Figure 2A). Notably, Level 1 contained the fewest different ontological terms (n=27) with the most representation in UMLS, and was therefore the least difficult task. Levels 2 and 3 were more complex (containing 102 and 150 distinct terms, respectively) and contained less representation in UMLS, as mentioned above (Table 3).

**Figure 2.**
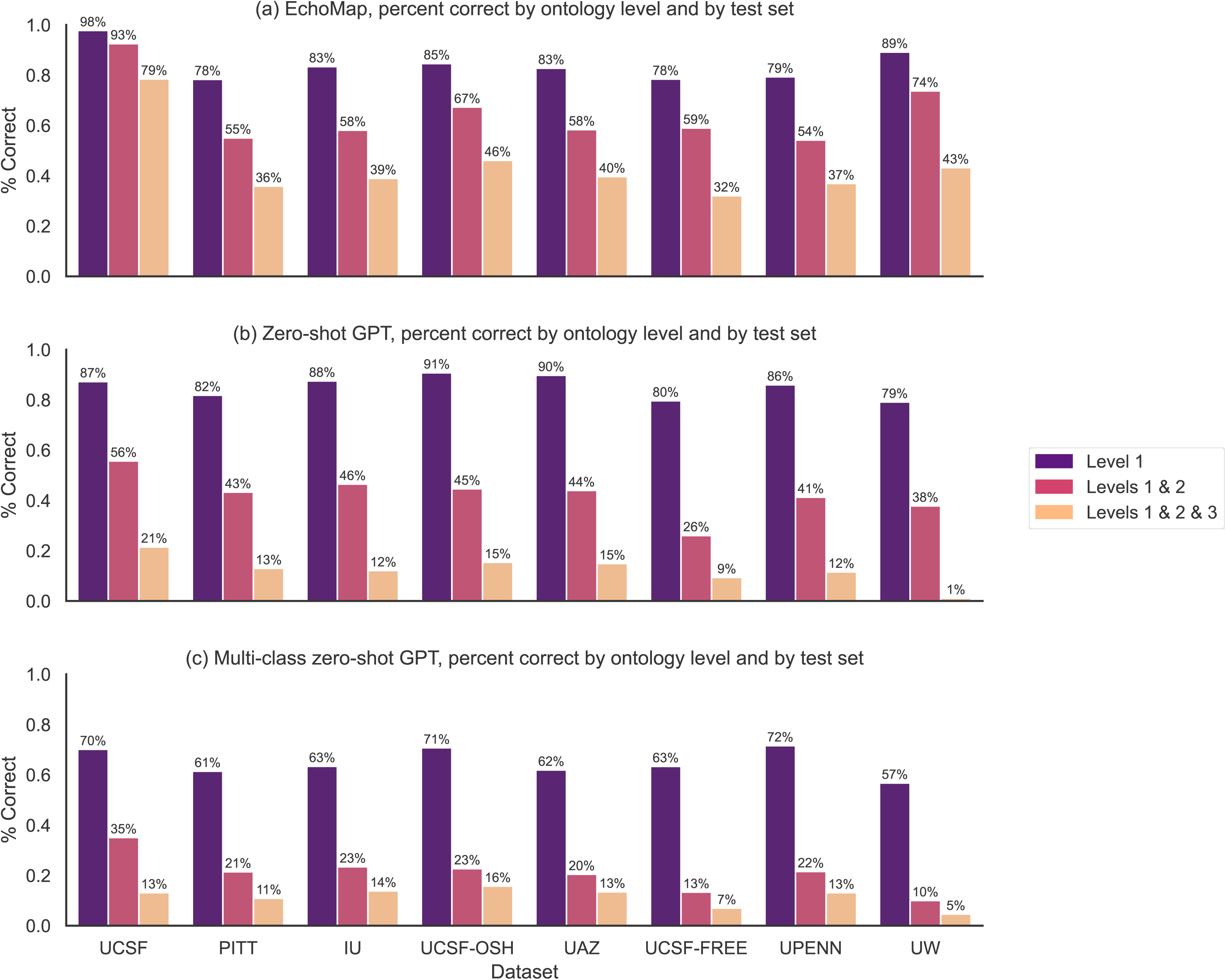
Correctness by ontology level, by dataset. Validation (UCSF) and test set (outside hospitals) performance for each of three model architectures: **(A)** Echomap, **(B)** Zero-shot GPT, **(C)** Multi-class zero-shot GPT. UCSF, University of California, San Francisco. PITT, University of Pittsburgh. IU, Indiana University. UCSF-OSH, outside hospital reports in the UCSF system. UAZ, University of Arizona. UCSF-FREE, free-text sentences from UCSF reports. UPENN, University of Pennsylvania. UW, University of Washington.

EchoMap retained good first-level performance across test datasets, with a mean of 82±4% correct on Level 1 (range 78-89%), 61±7% cumulatively on Levels 1 and 2(range 54%-74%), and 39±5% cumulatively on all three levels (range 32-46%, Figure 2A). Level 1 performance on the test datasets was lower than performance on the validation dataset (one-sample t-test p<0.001). For Levels 1 and 2 together, and for all three levels together, test dataset performance was also statistically significantly different from performance on the validation dataset (p<0.001, p<0.001 respectively).

### A small, statistical machine learning model was superior to single-shot inference from a large language model

Given the rapidly improving capabilities of newer LLMs for myriad language tasks, we also tested the ability of GPT to map echocardiogram text to the TTE ontology. Using one-shot inference allowed us to leverage the power of GPT without sacrificing any of the small echocardiogram dataset for training or fine-tuning; one-shot inference has recently been shown to meet or beat fine-tuned performance at some medical tasks^20^. GPT was used in two different ways. First, level 1, level 2, and level 3 were each predicted independently for each test sentence. Second, all three levels were predicted simultaneously for each sentence.

Using the independent classification approach, GPT gave 85±5% correct for Level 1 (range 79%-91%), 41±7% correct for Levels 1 and 2 (range 26%-46%), and 11±5% correct for all three levels (range 1%-15%). Using simultaneous classification for all three levels, performance was 64±5% for Level 1 (range 57%-72%), 19±5% for Levels 1 and 2 (range 10%-23%), and 11±4% for all three levels (range 5%-16%). These results are shown in Figure 2.

We compared test performance across the three methods: EchoMap, GPT with individual classifications, and multi-class GPT(Figure 1). EchoMap was statistically superior to GPT at all three ontology Levels (Friedman’s *X*^2^ p < 0.01 at Level1, p < 0.001 at Levels 1 and 2, and p < 0.01 at Levels 1, 2, and 3) (Figure 3). Only at Level 1, independent classification GPT, but not multi-class GPT, was statistically similar to EchoMap (pairwise p = 0.6 for individual GPT vs. EchoMap; p < 0.05 for multi-class GPT vs. EchoMap). At Levels 1, 2, and 3 together, both methods for GPT inference performed poorly and were statistically indistinguishable from each other (pairwise p = 0.9).

**Figure 3.**
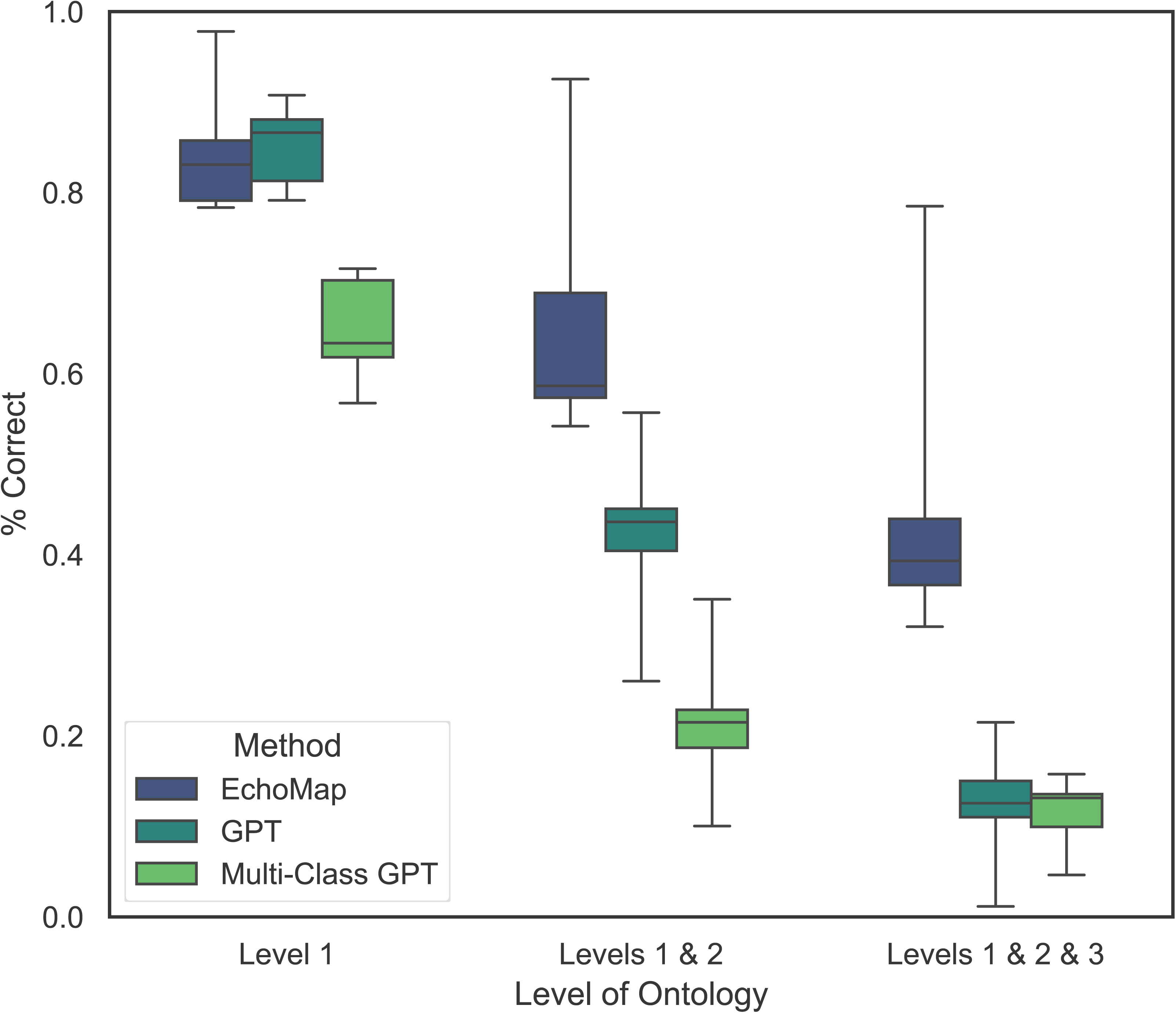
Aggregate performance across all datasets evaluated, by each mapping model and by ontology level. Box plots represent performance of all eight validation and test datasets in order to illustrate differences among mapping models.

### EchoMap performance relies most on UMLS entities and simple text overlap

In the development of the EchoMap statistical ML model, different engineered features were tested (see Methods). In addition to negation, Jaccard similarities, UMLS named entity recognition, and hierarchical predictions, all used in the final EchoMap model, position of speech embedding as well as BioWordVec embedding were also evaluated against the validation dataset for Level 1, Level 2, and Level 3 of the ontology (Supplemental Table 2).

Simple Jaccard indices between sentences and ontological terms worked well. Overall, UMLS named entity recognition was the most useful embedding compared to position of speech or BioWordVec. UMLS embeddings were more useful for L1 performance than for L2 or L3, likely consistent with the fact that fewer ontological terms from L2/L3 are currently represented in UMLS (Table 3).

### ML models displayed the ability to extrapolate to ontological combinations not initially included in training

Across all 7 external test sets, there were 2076 sentences where each level of Level 1, Level 2, and Level3 were able to be mapped using a rearrangement of existing ontology terms, even though that particular L1-L2-L3 combination was not present in the TTE ontology. EchoMap demonstrated an ability to map sentences not originally included in the training, mapping 359 of these sentences correctly. Multi-class GPT performed similarly, correctly mapping 354 of these sentences, while GPT making independent predictions per level performed worse, only mapping 128 correctly.

## Discussion

Harmonizing echocardiogram report text is critical to leverage it for cross-institutional big-data research. In this study, we provide proof of concept for how to apply NLP to harmonize echocardiogram report text.

Importantly, the models in this study do not just extract certain information from text but seek to map all echo report text to an ontology – a framework for knowledge. An ontology is key because, as Meta Chief AI Scientist Yann LeCun says, “there is a limit to how smart [language models] can be and how accurate they can be because they have no experience of the real world, which is really the underlying reality of language^21^.” The finding that eight different accredited hospitals operating according to echocardiography guidelines have such different data dictionaries is important in and of itself and demonstrates the need for greater harmonization.

One reason echocardiography report text has not been mapped to an ontology to date is the technical challenges inherent in doing so – challenges we address in this proof-of-concept study. Regular expressions-based text extraction is tedious and brittle, while the corpus of echo text is so small that ML is hard to implement – especially those large language models that seem to excel at more general language tasks. We show that a small statistical ML model, EchoMap, trained on a small amount of the echo report corpus can actually outperform one-shot inference from prevailing LLMs.

With EchoMap, we see a relationship between each level’s performance and the number of ontology terms present in UMLS; this suggests that further investment of adding echo-specific terms to UMLS (or alternatively, Radlex with greater software support for feature embeddings) could pay dividends both in improving EchoMap and in achieving greater representation of echocardiography concepts in more general medical ontologies.

### Limitations

There are several limitations of the current study, which we see as areas for future improvement.

A major limitation is that the echo ontology developed must be improved and expanded. The ontology used in the current study was created from only one institution’s data dictionary (along with clinical domain knowledge), so that other institutions’ dictionaries could be reserved for testing. An improved ontology could start with more data dictionaries, and could expand to all areas germane to echo reporting, including study quality, patient status, pediatric congenital heart disease, stress and transesophageal echocardiography, and more. With more institutional sharing of data dictionaries, the ontology can be improved.

Second, despite testing of several versions of EchoMap and two versions of GPT, more ML model exploration and improvement may further improve performance and generalizability.

### Conclusions

Mapping echocardiographic report text to a standardized ontology can aid in data mining efforts for both quality improvement and machine learning research. While LLMs have risen in popularity, small statistical machine learning models also perform well and may be especially useful for small, specialized text datasets where clinical meaning is important. These results highlight the utility of continuing to develop an open-source, high-resolution, standardized cardiac ontology to harmonize reports across institutions. One can envision a future where an optimized ontology is developed and maintained by the echocardiography community, with investment into depositing all terms into UMLS or similar general medical ontologies, and where all institutions can map their data dictionaries to this central and comprehensive resource.

## Data Availability

The UCSF data dictionary may be made available upon reasonable request for non-commercial use and with approval

## Abbreviations

(NLP): Natural language processing
(ML): machine learning
(LLM): large language model
(GPT): generative pre-trained transformer
(NER): named entity recognition

## Acknowledgements

We thank Marc Kohli and Ross Filice for background information about Radlex.

## Author Contributions

R.A. conceived of the manuscript. S.S., R.A., S.R., R.R., V.S. and B.G. performed analyses. R.A., S.S., H.A., J.T., R.T., E.A., R.M., and G.B.M. prepared and labeled data dictionaries and text. R.A., S.S. wrote the manuscript with input from all authors.

---

**Supplemental Table 1.** Extrapolation performance on term combinations outside TTE ontology.

|  | **Echomap** | **GPT** | **GPT Multi-Class** |
| --- | --- | --- | --- |
| **UCSF (n=0)** | n/a |  |  |
| **PITT (n=292)** |  |  |  |
| L1 | 237 (81%) | 260 (89%) | 199 (68%) |
| L1, L2 | 118 (40%) | 184 (63%) | 91 (31%) |
| L1, L2, L3 | 32 (11%) | 9 (3%) | 64 (22%) |
| **IU (n=321)** |  |  |  |
| L1 | 233 (73%) | 266 (83%) | 215 (67%) |
| L1, L2 | 137 (43%) | 186 (58%) | 71 (22%) |
| L1, L2, L3 | 46 (14%) | 22 (7%) | 45 (14%) |
| **UCSF-OSH (n=78)** |  |  |  |
| L1 | 41 (53%) | 61 (78%) | 38 (49%) |
| L1, L2 | 25 (32%) | 34 (44%) | 15 (19%) |
| L1, L2, L3 | 8 (10%) | 6 (8%) | 9 (12%) |
| **UAZ (n=268)** |  |  |  |
| L1 | 176 (66%) | 222 (83%) | 188 (70%) |
| L1, L2 | 90 (34%) | 131 (49%) | 62 (23%) |
| L1, L2, L3 | 46 (17%) | 35 (13%) | 46 (17%) |
| **UCSF-FREE (n=264)** |  |  |  |
| L1 | 152 (58%) | 177 (67%) | 137 (52%) |
| L1, L2 | 87 (33%) | 87 (33%) | 34 (13%) |
| L1, L2, L3 | 35 (13%) | 16 (6%) | 21 (8%) |
| **UPENN (n=667)** |  |  |  |
| L1 | 554 (83%) | 607 (91%) | 500 (75%) |
| L1, L2 | 272 (41%) | 407 (61%) | 267 (40%) |
| L1, L2, L3 | 149 (22%) | 40 (6%) | 160 (24%) |
| **UW (n=186)** |  |  |  |
| L1 | 136 (73%) | 112 (60%) | 82 (44%) |
| L1, L2 | 92 (49%) | 52 (28%) | 22 (12%) |
| L1, L2, L3 | 43 (23%) | 0 (0%) | 9 (5%) |

**Supplemental Table 2.** Iterations on statistical ML model features. Bold indicates the highest performance at each level. EchoMap is represented as the final entry in the table.

| Model Brief Description | Features used |  |  |  |  |  | Validation performance, % correct |  |  |
| --- | --- | --- | --- | --- | --- | --- | --- | --- | --- |
|  | Neg | Jac | Hier-archy | POS (PCA)* | BWV (PCA)* | UMLS (PCA)* | L1 | L2 | L3 |
| No Jaccard used | X |  | X | X (25) | X (25) | X (25) | 90 | 87 | 57 |
| Only embedding used is UMLS;<br>no Jaccard | X |  | X |  |  | X (25) | 91 | 83 | 54 |
| No embeddings used | X | X | X |  |  |  | 92 | 92 | 81 |
| Only embedding used is POS | X | X | X | X (25) |  |  | 95 | 90 | 80 |
| No UMLS embedding used | X | X | X | X (25) | X (25) |  | 95 | 92 | 81 |
| Only embedding used is BioWordVec | X | X | X |  | X (25) |  | 95 | 92 | 81 |
| Only embedding used is UMLS;<br>only 10 PCA elements | X | X | X |  |  | X (10) | 96 | 90 | **83** |
| No negation used |  | X | X | X (25) | X (25) | X (25) | 97 | 91 | 81 |
| No POS embedding used | X | X | X |  | X (25) | X (25) | 97 | 91 | 81 |
| UMLS, Jaccard, and Hierarchical only |  | X | X |  |  | X (25) | 97 | 92 | 80 |
| UMLS, and Jaccard only |  | X |  |  |  | X (25) | 97 | 92 | 80 |
| All features | X | X | X | X (25) | X (25) | X (25) | 97 | **93** | 82 |
| No BioWordVec embedding used;<br>no hierarchical pred | X | X |  | X (25) |  | X (25) | **98** | 92 | 79 |
| No BioWordVec embedding used | X | X | X | X (25) |  | X (25) | **98** | **93** | 81 |
| Only embedding used is UMLS;<br>no hierarchical pred | X | X |  |  |  | X (25) | **98** | **93** | 81 |
| Only embedding used is UMLS<br>(EchoMap) | X | X | X |  |  | X (25) | **98** | **93** | **83** |

\*indicates the number of principal components used
PCA, principal components analysis. Neg, negation detection. Jac, Jaccard indices. Hierarchy, Hierarchical predictions across levels. POS, position of speech tag embedding. BWV, BioWordVec embedding. UMLS, UMLS named entity recognition embedding.

## Figure Legends

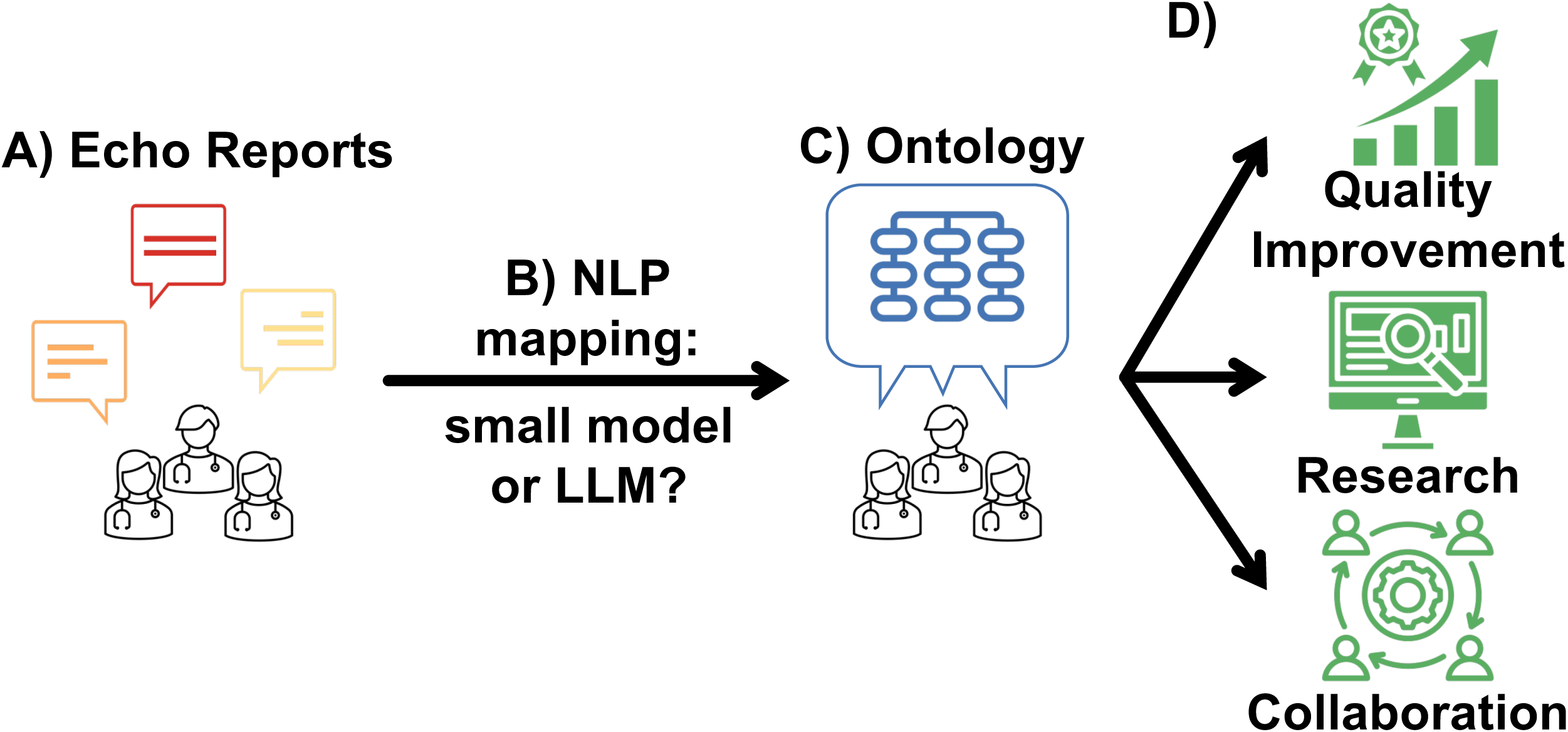
Central Illustration. **(A)** Echocardiography report text is a small, highly specialized corpus of text used to communicate echocardiography findings, and it includes both structured and free text that varies among institutions. **(B)** Mapping such text to a centralized ontology **(C)** for harmonization can improve quality improvement, big-data research, and cross-institution collaboration **(D)**. We tested the ability of natural language processing (NLP) to map echocardiography text to ontology, evaluating both a small statistical machine learning (ML) model as well as a large language model (LLM).

